# Inequalities in access to healthcare by local policy model among newly arrived refugees: evidence from population-based studies in two German states

**DOI:** 10.1101/2021.07.13.21260241

**Authors:** Judith Wenner, Louise Biddle, Nora Gottlieb, Kayvan Bozorgmehr

**Author notes:** **(corresponding author) Kayvan Bozorgmehr**.

## Abstract

**Background:** Access to healthcare is restricted for newly arriving asylum seekers and refugees (ASR) in many receiving countries, which may lead to inequalities in health. In Germany, regular access and full entitlement to healthcare (equivalent to statutory health insurance, SHI) is only granted after a waiting time of 18 months. During this time of restricted entitlements, local authorities implement different access models to regulate asylum seekers’ access to healthcare: the electronic health card (EHC) or the healthcare voucher (HV). This paper examines inequalities in access to healthcare by comparing healthcare utilization by ASR under the terms of different local models (i.e., regular access equivalent to SHI, EHC, and HV).

**Methods:** We used data from three population-based, cross-sectional surveys among newly arrived ASR (N=863) and analyzed six outcome measures: specialist and general practitioner (GP) utilization, unmet needs for specialist and GP services, emergency department use and avoidable hospitalization. Using logistic regression, we calculated odds ratios (OR) and 95% confidence intervals for all outcome measures, while considering need by adjusting for socio-demographic characteristics and health-related covariates.

**Results:** Compared to ASR with regular access, ASR under the HV model showed lower needs-adjusted odds of specialist utilization (OR=0.41 [0.24-0.66]) and a tendency towards lower GP (OR=0.61 [0.33-1.16]) and emergency department utilization (OR=0.74 [0.48-1.14]). ASR under the EHC model showed a tendency toward higher specialist unmet needs (OR= 1.89 [0.98-3.64]) and avoidable hospitalizations (OR=1.69 [0.87-3.30]) compared to ASR with regular access. A comparison between EHC and HV showed higher odds for specialist utilization under the EHC model as compared to the HV model (OR=2.39 [1.03-5.52]).

**Conclusion:** ASR using the HV are disadvantaged in their access to healthcare compared to ASR having either an EHC or regular access. Given equal need, they use specialist (and partly also GP) services less. The identified inequalities constitute inequities in access to healthcare that could be reduced by policy change from HV to the EHC model during the initial 18 months waiting time, or by granting ASR regular healthcare access upon arrival. Minor differences in unmet needs, emergency department use and avoidable hospitalization between the models deserve further exploration in future studies.

## Background

Providing access to healthcare for asylum seekers and refugees (ASR) is part of the receiving countries’ legal responsibilities [1]. However, many countries restrict the access of newly arriving ASR to regular healthcare services [2]. This may hinder need-based healthcare utilization, impact the health of ASR, and lead to inequalities in health [3, 4]. Monitoring the access to healthcare of ASR is therefore an important public health task.

There are multiple ways of looking at access. Following Aday and Andersen [5], access to healthcare means that those “who need care get into the system” (p. 218). It can be measured as *potential access* [6], with a focus on availability of services and insurance coverage. However, this tells us little about the actual utilization and ignores the impact of social determinants beyond health system characteristics on healthcare utilization [7, 8]. Alternatively, access can be equated with *realized access* [6]. It can then be conceptualized as actual use of healthcare services (using utilization indicators) or as non-realized access (using outcome indicators like forgone care and unmet needs) [9]. Further measures include ambulatory care sensitive hospitalization, which measures the lack of timely provision of ambulatory treatment leading to a potentially avoidable hospitalization [10].

When looking at determinants of realized access to healthcare among newly arriving ASR, the legal framework must be considered. In Germany, newly arriving asylum seekers are excluded from statutory health insurance (SHI). Their health entitlements are regulated by the asylum seekers’ benefits act (in German: Asylbewerberleistungsgesetz (AsylbLG)), which is a federal law. Compared to SHI, the AsylbLG grants only a limited scope of healthcare during the first 18 months (15 months at time of data collection) or until a permanent protection status (refugee status or subsidiary protection) has been granted. The restricted entitlements include healthcare in case of acute illnesses and pain, preventive services and vaccines, and services related to pregnancy and birth (AsylbLG Art. 4). Access to further, mostly specialized, services can be granted on a case-by-case basis (AsylbLG Art. 6).

Given Germany’s federal structure and decentralized governance, the local governments and social welfare offices (SWO) are responsible for implementing the AsylbLG, and thus for organizing access to healthcare for ASR [11]. Different policy choices on state and local levels have led to two different access models being applied across Germany to implement the AsylbLG during the waiting period: the health care voucher (in the following: HV) and the electronic health card (in the following: EHC) [12, 13].

After completing the 18 months waiting period, the AsylbLG entitles asylum seekers to a scope of healthcare that is equivalent to SHI (AsylbLG Art. 2). Upon obtaining permanent legal status, refugees have access to healthcare via SHI membership [11]. In both cases, they use a health insurance card to access health services. Hence, in practice, both recognized refugees and asylum seekers after 18 months in Germany have regular access, i.e. SHI- or SHI-equivalent entitlements to healthcare. For the purpose of our study, we thus distinguish between three different access models:

1. **Health care vouchers** (**HVs**), which allow for healthcare access during the first 18 months in Germany. HVs are issued by the local SWO and entitlements are restricted (AsylbLG Art. 4 and 6). The paper-based HVs are usually valid for three months, or a single visit to a healthcare provider, and to be used within the respective administrative district. They are deposited with one service provider per calendar quarter, and each referral necessitates the approval and dispensation of another HV by the local SWO [11].
2. **Electronic health cards** (**EHCs**), which allow for healthcare access during the first 18 months in Germany. The EHCs are issued by a local statutory health insurance (SHI) fund, but financed by the SWOs. Entitlements are restricted (AsylbLG Art. 4 and 6). The EHC is issued once and then usually valid for the whole period of restricted entitlements. It has a digital record of patient details and stays with the patient. Though EHC holders do not become members of the SHI, the SHI carries out billing and accounting procedures against an administrative fee.
3. Irrespective of the access model used during the 18 months waiting period (HV or EHC), restrictions on healthcare entitlements and access are lifted after 18 months by granting SHI-equivalent health benefits (regulated by the AsylbLG Art. 2 and financed through the SWOs); or earlier if full SHI membership is granted through a temporary or permanent residence permit (**regular access**) [13–16].

The choice of access model during the first 18 months – HV oder EHC – has been subject to controversial political debates. Proponents of the EHC model claim that it reduces discrimination against ASR, facilitates need-based healthcare utilization and reduces the administrative workload for welfare offices and service providers. Proponents of the HV model caution that the EHC model will lead to excessive healthcare utilization and thereby increase expenses [13, 17, 18].

So far, there is limited evidence on the impact of the local policy model on access to healthcare. Qualitative studies suggest that the HVs are difficult to handle for healthcare users and providers and thereby hamper access to health services [18–21]. Quantitative studies provide further evidence for the disadvantages of the HV and of the entitlement restrictions during the 18-month waiting period; for instance, in terms of higher medical costs [22–25]. So far, there is no quantitative evidence of inequalities in access to healthcare among ASR who are subject to the three different access models. The aim of our research was therefore to analyze if different access models (HV, EHC, regular access) are associated with inequalities in access to healthcare understood as realized access or forgone care.

## Methods

### Design, sampling, and population

We used data from three population-based, cross-sectional surveys among newly arrived ASR (N=863) living in accommodation or reception centres in the states of Baden-Württemberg (BW) and Berlin (BE). In BE, the EHC was introduced in 2016; whereas in BW, all municipalities use HVs [11]. Sampling, recruitment, and survey instruments were nearly identical in both states [26, 27]. Around 3% of all 2,017 accommodation centres across the two states (n=81) were selected using random sampling and all adult residents of these centres (census approach) were invited to participate in the survey. In addition, six reception centres from BW were purposively selected for inclusion, with 25% of residents selected by random sampling and invited to participate. The overall response rate was 30.5% (see additional file 1). Questionnaires were developed from standardised, international survey instruments. They covered health status, access to and utilization of health services and socio-demographic aspects. Participants filled out a paper questionnaire in one of nine languages. Data collection for the majority of respondents (96% of the sample) took place between January 2018 and November 2018, while less than 4% of participants (32 persons) were recruited in December 2019. The study design, sampling procedure and data collection process have been described in more detail elsewhere [26, 27].

### Outcome measures

A wide range of utilization (or process) indicators have been suggested to measure realized access [6, 7]. While utilization indicators are important to detect barriers and assess equity in access to health services, utilization is not always an aim in itself [8]. Therefore, in addition to utilization indicators, outcome indicators related to the health consequences of service utilization (vs. forgone or delayed care) are also commonly included in the measurement of access. Subjective unmet needs and avoidable hospitalizations are two important outcome indicators that are internationally used to this end [10, 28]. They have been adapted to the German context [29, 30] and to refugee populations in Germany in particular [31–33]. Subjective unmet need describes a situation in which healthcare was not sought despite subjectively felt need [28]. Avoidable hospitalization can be defined as hospital admissions for conditions for which hospitalization can be prevented by providing timely and adequate treatment in the outpatient setting. These conditions are defined as ambulatory care-sensitive conditions (ACSC) [10].

To analyse differences in access to healthcare, we included three utilization and three outcome indicators. As utilization indicators we included self-reported utilization of general practitioner (GP) and specialist services in the last four weeks (y/n) and of emergency departments in the last 12 months (y/n). As outcome indicators we included hospital admissions for ACSC in the last 12 months (y/n) and subjective unmet needs for specialist or GP services in the last 12 months (y/n). ACSC were assessed using two questions: first, participants were asked whether they had one of the conditions identified as ACSC by Sundmacher et al. [29] in the last 12 months (see additional file 2). Second, they were asked whether they had been hospitalized for any of the said conditions. To assess unmet needs, participants were asked directly if they had refrained from seeking healthcare despite the subjectively felt need to see a doctor. Results for the latter question were only available for participants from BW.

### Exposures and Co-variables

The access model used – HV, EHC or regular access – was set as the exposure. It was directly assessed for the state of BW. For Berlin, all persons with a duration of stay of more than 15 months or with a secured residence status (refugee status or subsidiary status) were coded as having regular access, while all others were considered using an EHC. It is important to note that this “waiting period” was extended from 15 to 18 months in August 2019. That is, at the time of data collection, restricted health entitlements applied during asylum seekers’ first 15 months in Germany. For this reason, we used a duration of stay of 15 months as a cut-off date in our data analysis.

An approach to differentiate among determinants of inequalities in realized access has been developed by Andersen and colleagues. They distinguish between predisposing characteristics (e.g., age, sex, socioeconomic status) and enabling resources which are related to the health system (e.g., health policy, financing, organization, resources, and availability of services). Given the importance of need as a major determinant of healthcare utilization, considering the actual health status – and thereby approximating healthcare needs – is essential [5, 6, 34]. We therefore included major predisposing characteristics (age, sex, region of origin, duration of stay in Germany, accommodation type and education) and important need- and health-related information (subjective health, chronic illness, and having a regular GP) as covariates.

### Statistical Analysis

We used logistic regression to calculate odds ratios (OR) for all outcomes, adjusting for socio-demographic characteristics (age, sex, region of origin, duration of stay in Germany, accommodation type and education) and health-related covariates (subjective health, chronic illness and having a regular GP) which we identified as potential confounders. Having regular access – as compared to access via EHC or HV – was chosen as a reference category. Based on the literature, this was considered the best possible access option. In addition, the HV model was also used as reference category repeating the same regression analyses for all outcomes. This allowed for a direct comparison of differences in access between the HV and EHC model.

All analyses were weighted (using design and calibration weights), treating reception centres in BW, accommodation centres in BW, and accommodation centres in BE each as separate clusters (see additional file 3). Calibration was conducted using data from the statistical offices in BW and BE for age, sex and region of origin [35, 36]. Missing values did not show systematic patterns related to the outcome and were thus assumed to be missing at random. For outcome and exposure indicators, missing values were imputed using single imputation according to the R-package *mice* [37] (see additional file 4). To understand the sensitivity of our results to weighting, the design effect (DEFF) was calculated. Low DEFF indicate small weighting effects. The overall model fit comparing the differences between observed and expected values for the Null-model and the full model was assessed using an adapted F-test for weighted survey designs. Larger F-values with non-significant p-values (>0.05) indicate better model fit [38].

## Results

### Descriptive results

The sample includes responses from 863 individuals of which 560 were living in the state of BW and 303 in the state of BE. Of the 560 participants in BW, 250 (44.6%) were using the HV model and 240 (42.7%) reported regular access. For 70 individuals (12.5%), information on the access model was missing. In Berlin, 49 (16.2%) were using the EHC model and 227 (74.9%) were having regular access while information on the access model was missing for 27 participants (8.9%).

There were no significant differences in age, sex, educational score or health status (subjective health or chronic illnesses) between persons subject to different access models. Given the requirements for regular access (either duration of stay of more than 15 months at the time of data collection or refugee status), duration of stay and residence status are highly associated with the access model. The region of origin is also significantly associated with the access model (c. Tab. 1 for details).

[Table 1]

**Table 1:**
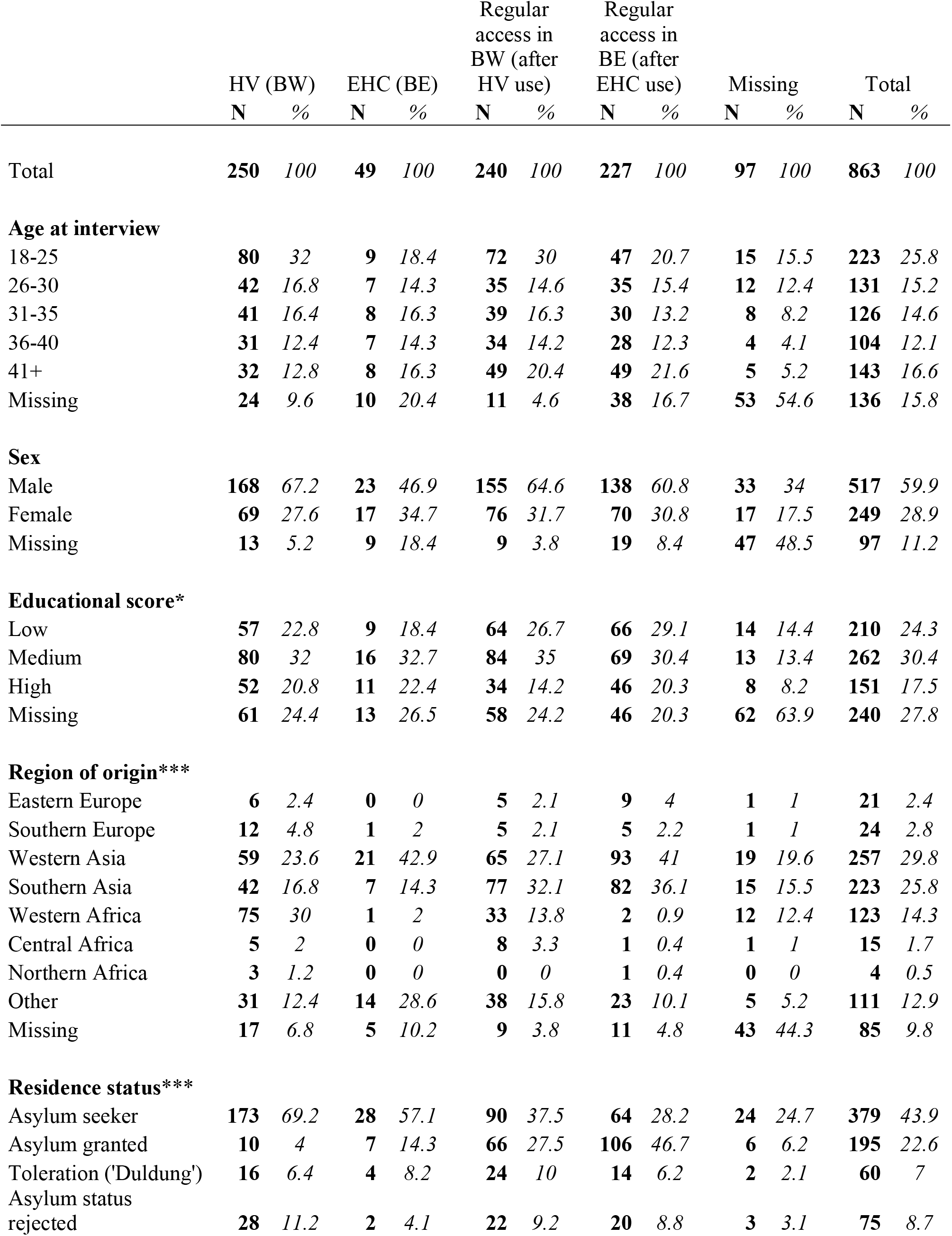
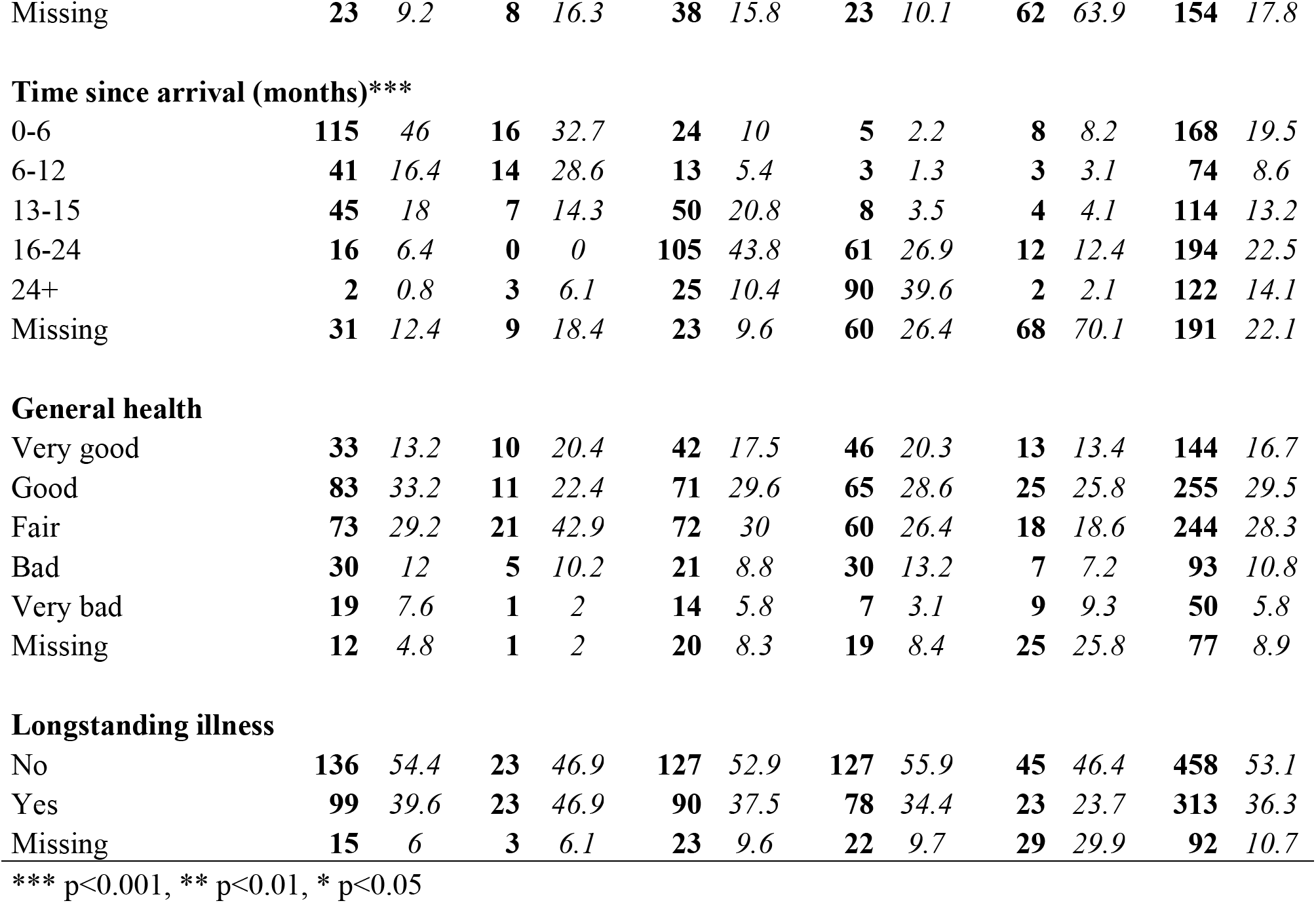
Socio-demographic and health-related information of the sample according to access model.

|  | HV (BW) |  | EHC (BE) |  | Regular access in BW (after HV use) |  | Regular access in BE (after EHC use) |  | Missing |  | Total |  |
| --- | --- | --- | --- | --- | --- | --- | --- | --- | --- | --- | --- | --- |
|  | N | % | N | % | N | % | N | % | N | % | N | % |
| Total | 250 | 100 | 49 | 100 | 240 | 100 | 227 | 100 | 97 | 100 | 863 | 100 |
| **Age at interview** |  |  |  |  |  |  |  |  |  |  |  |  |
| 18-25 | 80 | 32 | 9 | 18.4 | 72 | 30 | 47 | 20.7 | 15 | 15.5 | 223 | 25.8 |
| 26-30 | 42 | 16.8 | 7 | 14.3 | 35 | 14.6 | 35 | 15.4 | 12 | 12.4 | 131 | 15.2 |
| 31-35 | 41 | 16.4 | 8 | 16.3 | 39 | 16.3 | 30 | 13.2 | 8 | 8.2 | 126 | 14.6 |
| 36-40 | 31 | 12.4 | 7 | 14.3 | 34 | 14.2 | 28 | 12.3 | 4 | 4.1 | 104 | 12.1 |
| 41+ | 32 | 12.8 | 8 | 16.3 | 49 | 20.4 | 49 | 21.6 | 5 | 5.2 | 143 | 16.6 |
| Missing | 24 | 9.6 | 10 | 20.4 | 11 | 4.6 | 38 | 16.7 | 53 | 54.6 | 136 | 15.8 |
| **Sex** |  |  |  |  |  |  |  |  |  |  |  |  |
| Male | 168 | 67.2 | 23 | 46.9 | 155 | 64.6 | 138 | 60.8 | 33 | 34 | 517 | 59.9 |
| Female | 69 | 27.6 | 17 | 34.7 | 76 | 31.7 | 70 | 30.8 | 17 | 17.5 | 249 | 28.9 |
| Missing | 13 | 5.2 | 9 | 18.4 | 9 | 3.8 | 19 | 8.4 | 47 | 48.5 | 97 | 11.2 |
| **Educational score\*** |  |  |  |  |  |  |  |  |  |  |  |  |
| Low | 57 | 22.8 | 9 | 18.4 | 64 | 26.7 | 66 | 29.1 | 14 | 14.4 | 210 | 24.3 |
| Medium | 80 | 32 | 16 | 32.7 | 84 | 35 | 69 | 30.4 | 13 | 13.4 | 262 | 30.4 |
| High | 52 | 20.8 | 11 | 22.4 | 34 | 14.2 | 46 | 20.3 | 8 | 8.2 | 151 | 17.5 |
| Missing | 61 | 24.4 | 13 | 26.5 | 58 | 24.2 | 46 | 20.3 | 62 | 63.9 | 240 | 27.8 |
| **Region of origin\*\*\*** |  |  |  |  |  |  |  |  |  |  |  |  |
| Eastern Europe | 6 | 2.4 | 0 | 0 | 5 | 2.1 | 9 | 4 | 1 | 1 | 21 | 2.4 |
| Southern Europe | 12 | 4.8 | 1 | 2 | 5 | 2.1 | 5 | 2.2 | 1 | 1 | 24 | 2.8 |
| Western Asia | 59 | 23.6 | 21 | 42.9 | 65 | 27.1 | 93 | 41 | 19 | 19.6 | 257 | 29.8 |
| Southern Asia | 42 | 16.8 | 7 | 14.3 | 77 | 32.1 | 82 | 36.1 | 15 | 15.5 | 223 | 25.8 |
| Western Africa | 75 | 30 | 1 | 2 | 33 | 13.8 | 2 | 0.9 | 12 | 12.4 | 123 | 14.3 |
| Central Africa | 5 | 2 | 0 | 0 | 8 | 3.3 | 1 | 0.4 | 1 | 1 | 15 | 1.7 |
| Northern Africa | 3 | 1.2 | 0 | 0 | 0 | 0 | 1 | 0.4 | 0 | 0 | 4 | 0.5 |
| Other | 31 | 12.4 | 14 | 28.6 | 38 | 15.8 | 23 | 10.1 | 5 | 5.2 | 111 | 12.9 |
| Missing | 17 | 6.8 | 5 | 10.2 | 9 | 3.8 | 11 | 4.8 | 43 | 44.3 | 85 | 9.8 |
| **Residence status\*\*\*** |  |  |  |  |  |  |  |  |  |  |  |  |
| Asylum seeker | 173 | 69.2 | 28 | 57.1 | 90 | 37.5 | 64 | 28.2 | 24 | 24.7 | 379 | 43.9 |
| Asylum granted | 10 | 4 | 7 | 14.3 | 66 | 27.5 | 106 | 46.7 | 6 | 6.2 | 195 | 22.6 |
| Toleration ('Duldung') | 16 | 6.4 | 4 | 8.2 | 24 | 10 | 14 | 6.2 | 2 | 2.1 | 60 | 7 |
| Asylum status rejected | 28 | 11.2 | 2 | 4.1 | 22 | 9.2 | 20 | 8.8 | 3 | 3.1 | 75 | 8.7 |
| Missing | **23** | *9.2* | **8** | *16.3* | **38** | *15.8* | **23** | *10.1* | **62** | *63.9* | **154** | *17.8* |
| **Time since arrival (months)\*\*\*** |  |  |  |  |  |  |  |  |  |  |  |  |
| 0-6 | **115** | *46* | **16** | *32.7* | **24** | *10* | **5** | *2.2* | **8** | *8.2* | **168** | *19.5* |
| 6-12 | **41** | *16.4* | **14** | *28.6* | **13** | *5.4* | **3** | *1.3* | **3** | *3.1* | **74** | *8.6* |
| 13-15 | **45** | *18* | **7** | *14.3* | **50** | *20.8* | **8** | *3.5* | **4** | *4.1* | **114** | *13.2* |
| 16-24 | **16** | *6.4* | **0** | *0* | **105** | *43.8* | **61** | *26.9* | **12** | *12.4* | **194** | *22.5* |
| 24+ | **2** | *0.8* | **3** | *6.1* | **25** | *10.4* | **90** | *39.6* | **2** | *2.1* | **122** | *14.1* |
| Missing | **31** | *12.4* | **9** | *18.4* | **23** | *9.6* | **60** | *26.4* | **68** | *70.1* | **191** | *22.1* |
| **General health** |  |  |  |  |  |  |  |  |  |  |  |  |
| Very good | **33** | *13.2* | **10** | *20.4* | **42** | *17.5* | **46** | *20.3* | **13** | *13.4* | **144** | *16.7* |
| Good | **83** | *33.2* | **11** | *22.4* | **71** | *29.6* | **65** | *28.6* | **25** | *25.8* | **255** | *29.5* |
| Fair | **73** | *29.2* | **21** | *42.9* | **72** | *30* | **60** | *26.4* | **18** | *18.6* | **244** | *28.3* |
| Bad | **30** | *12* | **5** | *10.2* | **21** | *8.8* | **30** | *13.2* | **7** | *7.2* | **93** | *10.8* |
| Very bad | **19** | *7.6* | **1** | *2* | **14** | *5.8* | **7** | *3.1* | **9** | *9.3* | **50** | *5.8* |
| Missing | **12** | *4.8* | **1** | *2* | **20** | *8.3* | **19** | *8.4* | **25** | *25.8* | **77** | *8.9* |
| **Longstanding illness** |  |  |  |  |  |  |  |  |  |  |  |  |
| No | **136** | *54.4* | **23** | *46.9* | **127** | *52.9* | **127** | *55.9* | **45** | *46.4* | **458** | *53.1* |
| Yes | **99** | *39.6* | **23** | *46.9* | **90** | *37.5* | **78** | *34.4* | **23** | *23.7* | **313** | *36.3* |
| Missing | **15** | *6* | **3** | *6.1* | **23** | *9.6* | **22** | *9.7* | **29** | *29.9* | **92** | *10.7* |

\*\*\* p<0.001, \*\* p<0.01, \* p<0.05

Of all participants, 29% indicated having used a specialist and 43.3% reported having seen a GP in the last four weeks. There was a considerable difference between the states for specialist utilization with only 24.1% reporting a visit to a specialist in BW compared to 38% in BE. We found no substantial differences in emergency department use, subjective unmet needs of specialist and GP services, and avoidable hospitalization. In total, 27.9% reported at least one visit to the emergency department, 26% and 26.7% reported unmet needs for specialist and GP services respectively, and 21.3% reported at least one avoidable hospitalization in the last 12 months. The share of missing information was rather high for specialist utilization (24.1%), but also for GP utilization (18.1%) and unmet needs (19.7% and 18.7%), while it was only 10.7% for emergency department use and 7.1% for avoidable hospitalization (c. Tab. 2).

**Table 2:**
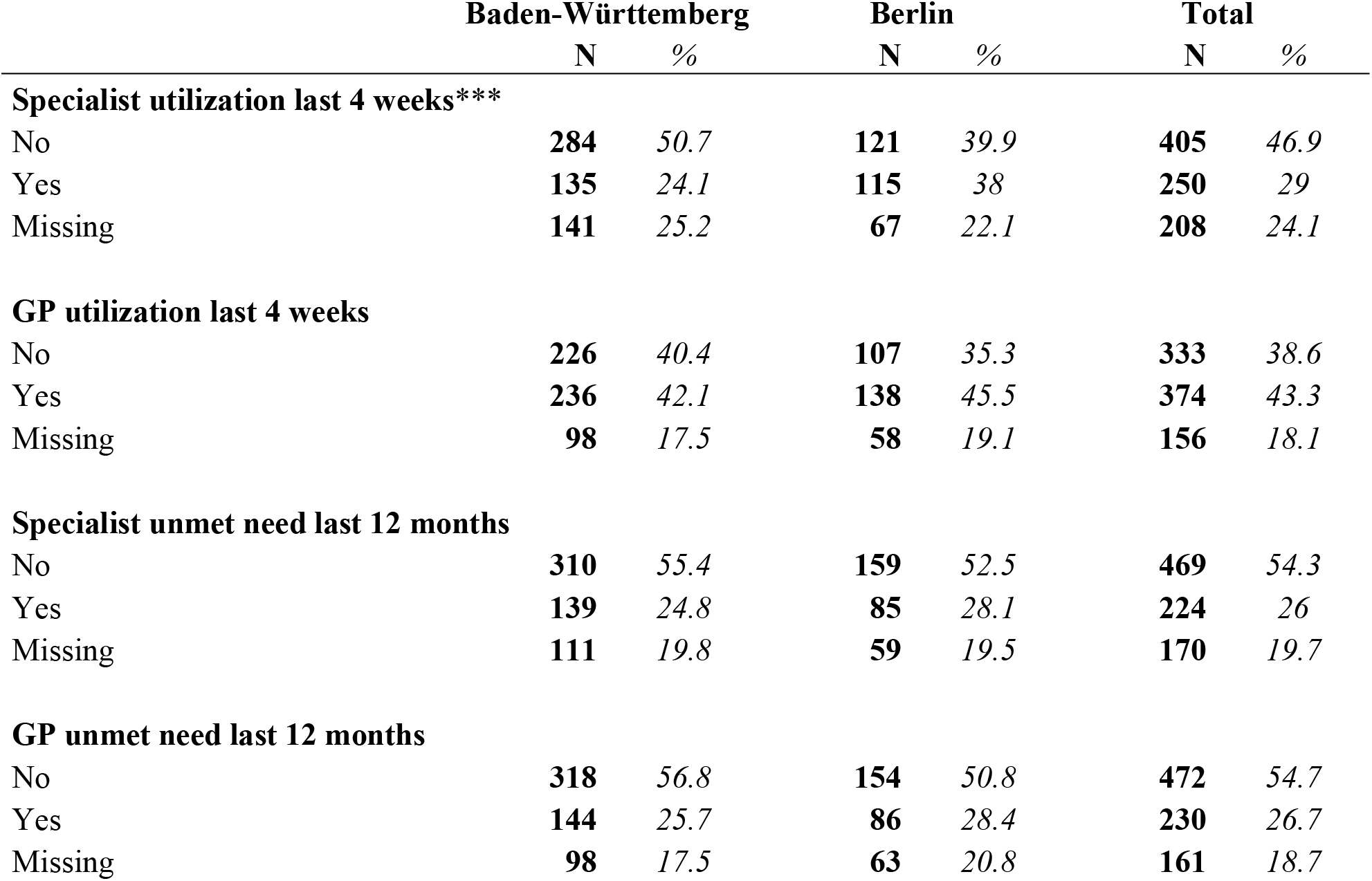
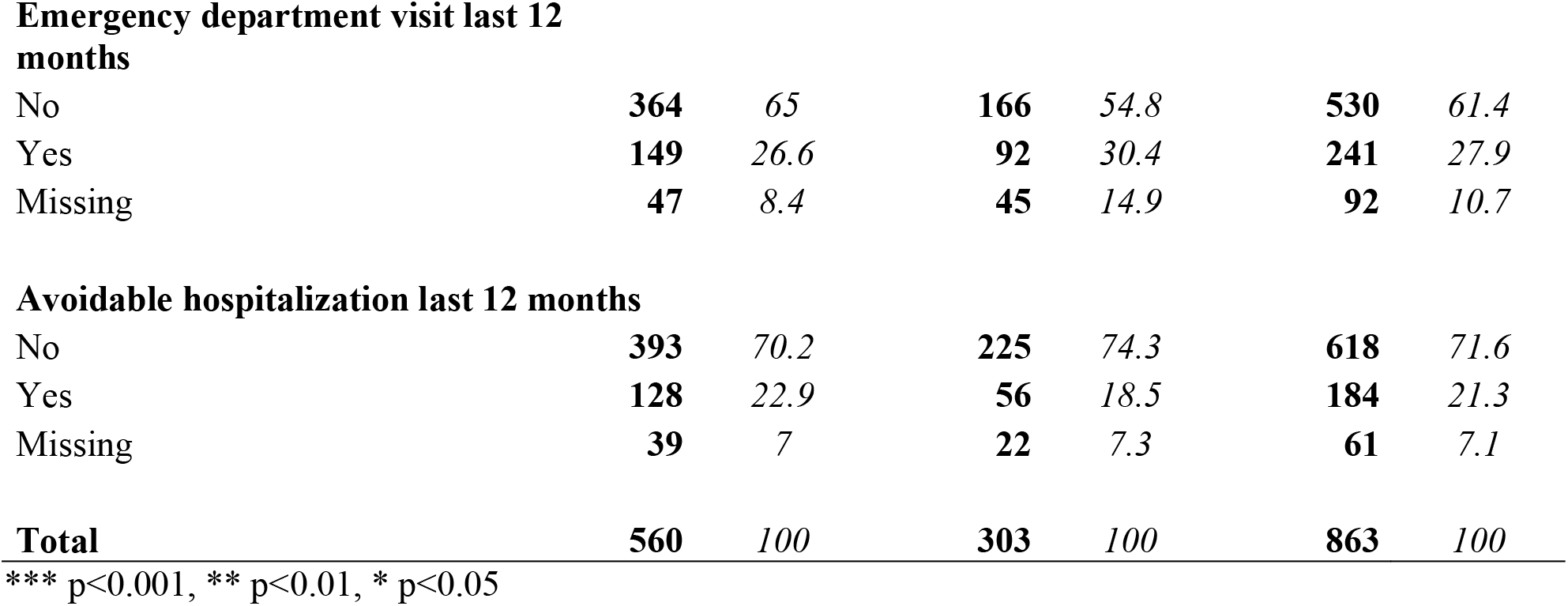
Utilization and outcome measures according to state.

|  | Baden-Württemberg |  | Berlin |  | Total |  |
| --- | --- | --- | --- | --- | --- | --- |
|  | N | % | N | % | N | % |
| **Specialist utilization last 4 weeks\*\*\*** |  |  |  |  |  |  |
| No | **284** | *50.7* | **121** | *39.9* | **405** | *46.9* |
| Yes | **135** | *24.1* | **115** | *38* | **250** | *29* |
| Missing | **141** | *25.2* | **67** | *22.1* | **208** | *24.1* |
| **GP utilization last 4 weeks** |  |  |  |  |  |  |
| No | **226** | *40.4* | **107** | *35.3* | **333** | *38.6* |
| Yes | **236** | *42.1* | **138** | *45.5* | **374** | *43.3* |
| Missing | **98** | *17.5* | **58** | *19.1* | **156** | *18.1* |
| **Specialist unmet need last 12 months** |  |  |  |  |  |  |
| No | **310** | *55.4* | **159** | *52.5* | **469** | *54.3* |
| Yes | **139** | *24.8* | **85** | *28.1* | **224** | *26* |
| Missing | **111** | *19.8* | **59** | *19.5* | **170** | *19.7* |
| **GP unmet need last 12 months** |  |  |  |  |  |  |
| No | **318** | *56.8* | **154** | *50.8* | **472** | *54.7* |
| Yes | **144** | *25.7* | **86** | *28.4* | **230** | *26.7* |
| Missing | **98** | *17.5* | **63** | *20.8* | **161** | *18.7* |
| **Emergency department visit last 12 months** |  |  |  |  |  |  |
| No | **364** | *65* | **166** | *54.8* | **530** | *61.4* |
| Yes | **149** | *26.6* | **92** | *30.4* | **241** | *27.9* |
| Missing | **47** | *8.4* | **45** | *14.9* | **92** | *10.7* |
| **Avoidable hospitalization last 12 months** |  |  |  |  |  |  |
| No | **393** | *70.2* | **225** | *74.3* | **618** | *71.6* |
| Yes | **128** | *22.9* | **56** | *18.5* | **184** | *21.3* |
| Missing | **39** | *7* | **22** | *7.3* | **61** | *7.1* |
| **Total** | **560** | *100* | **303** | *100* | **863** | *100* |

\*\*\* p<0.001, \*\* p<0.01, \* p<0.05

### Inequalities in access comparing HV and EHC with regular access

ASR under the HV model were less likely to use specialist (OR=0.46 [0.31-0.70]), GP (OR=0.57 [0.34-0.95]) and emergency department services (OR=0.76 [0.51-1.14]) compared to ASR with regular access while adjusting for age and sex. For specialist and GP utilization this difference can be considered significant based on the 95%-CI. Outcome indicators did not differ significantly between both groups.

ASR under the EHC model were more likely to report unmet needs for specialist services (OR=2.11 [1.32-3.40]) and showed a tendency toward higher avoidable hospitalization (OR=1.51 [0.81-2.81]) compared to ASR with regular access while adjusting for age and sex. Point estimates for specialist and GP utilization showed lower but non-significant ORs among ASR with EHC compared to regular access, while no considerable difference was found for both GP unmet needs and emergency department visits between the two groups (c. Fig. 1).

**Figure 1:**
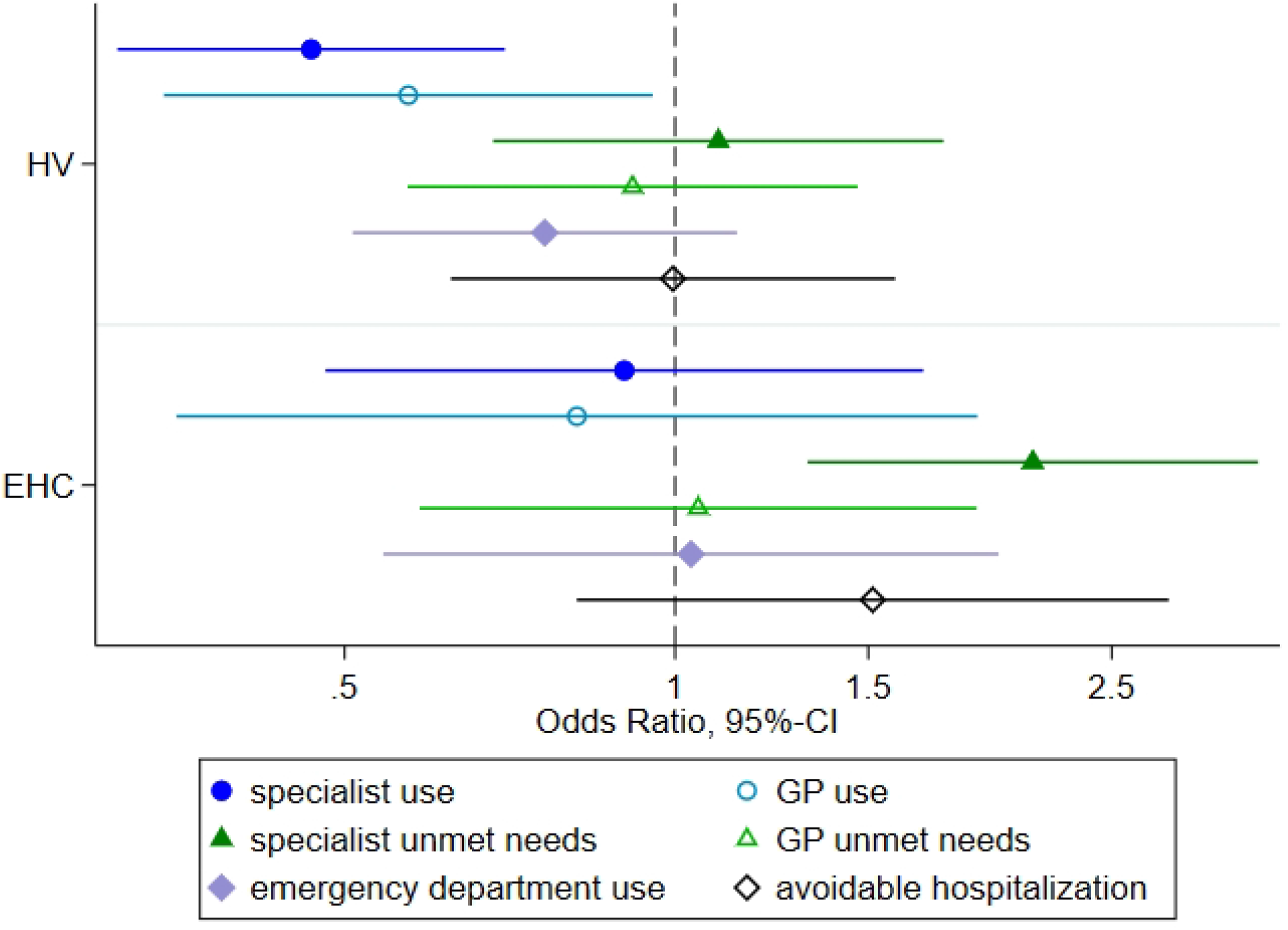
Odds-Ratios (and 95%-CIs) of access to healthcare comparing between access models, adjusted for age and sex (ref=regular access) Legend: HV=Health care voucher; EHC=electronic health card; x-axis with 95% confidence intervals on a log-scale

The final models were adjusted for health status, duration of stay, region of origin, educational score, having a regular GP and accommodation type. Their results were similar to those of the simple models. ASR under the HV model showed lower odds of specialist (OR=0.41 [0.24-0.66]) and a tendency towards lower GP utilization (OR=0.61 [0.33-1.16]) and emergency department utilization (OR=0.74 [0.48-1.14]) compared to ASR with regular access. For all other indicators, there was no difference between HV users and ASR with regular access.

ASR under the EHC model showed a tendency toward higher specialist unmet needs (OR= 1.89 [0.98-3.64]) and avoidable hospitalizations (OR=1.69 [0.87-3.30]) compared to ASR with regular access in the fully adjusted models. For all other outcomes, the differences between ASR under the EHC model and regular access were negligible (c. Fig. 2).

**Figure 2:**
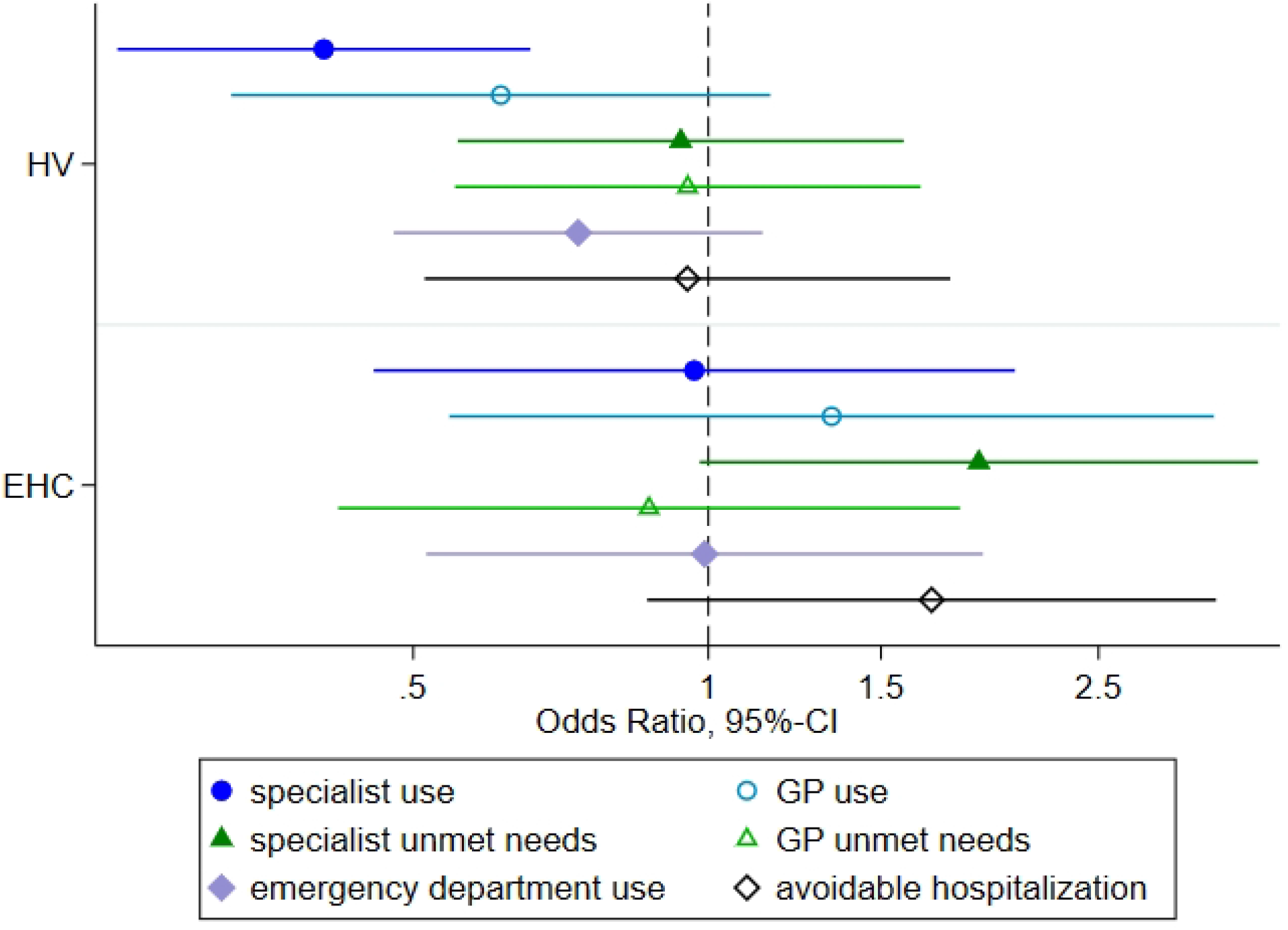
Fully adjusted Odds-Ratios (and 95%-CIs) of access to healthcare comparing between access models (ref=regular access) Legend: HV=Health care voucher; EHC=electronic health card; x-axis with 95% confidence intervals on a log-scale

### Inequalities in access comparing the HV and the EHC model

The comparison of access between the two models used during the 15 months waiting period showed higher odds for specialist utilization (OR=1.93 [1.01-3.69]) and specialist unmet needs (OR=1.94 [1.13-3.31]) among ASR under the EHC model compared to ASR under the HV model, adjusting for age and sex. For the remaining four outcomes, odds among ASR with a EHC were also higher compared to odds among ASR with HVs. However, given wide confidence intervals that include the value one, they do not indicate significant differences (c. Fig. 3).

**Figure 3:**
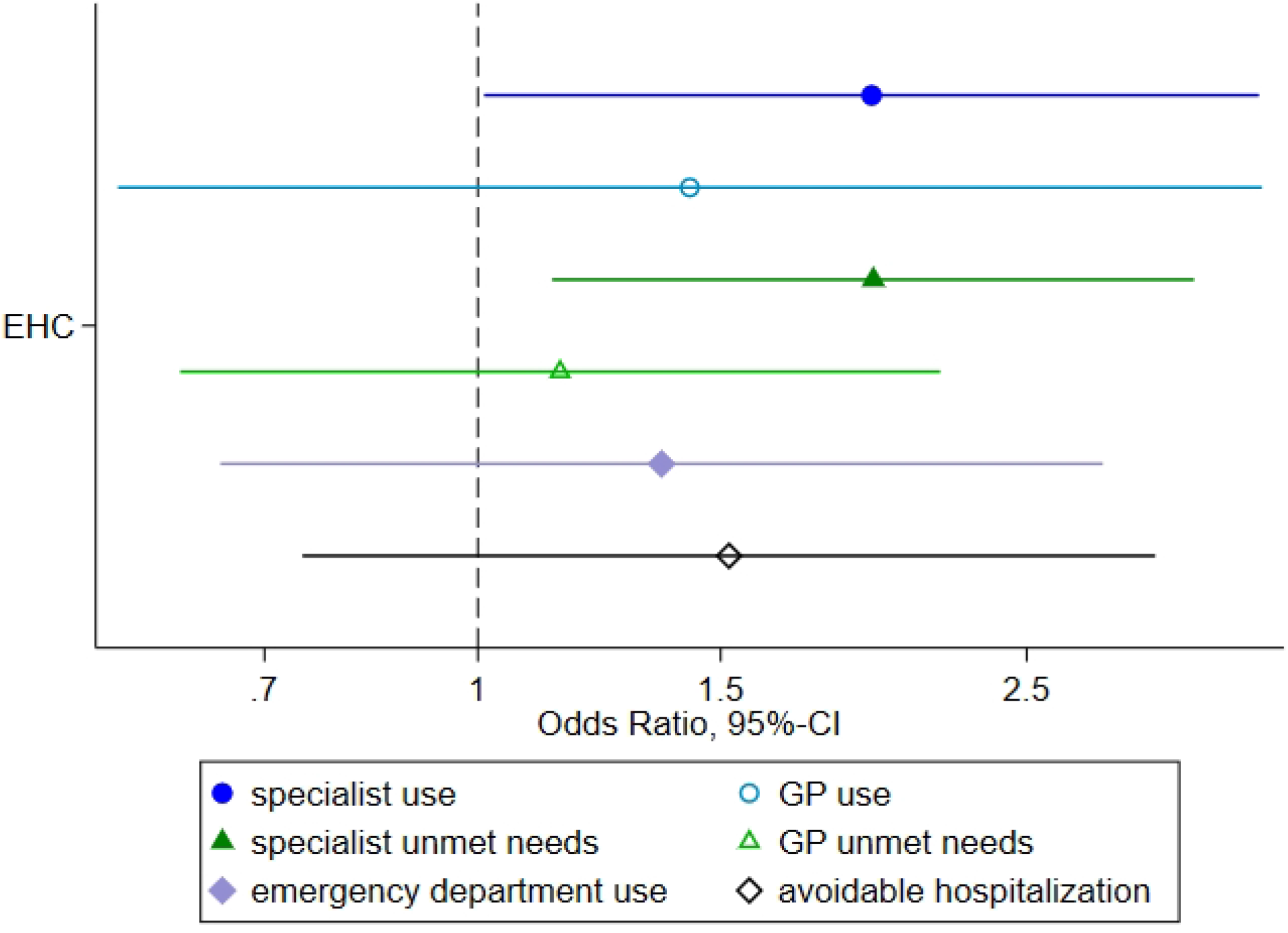
Odds-Ratios (and 95%-CIs) of access to healthcare comparing between access models used in the first 15 months, adjusted for age and sex (ref=HV) Legend: HV=Health care voucher; EHC=electronic health card; x-axis with 95% confidence intervals on a log-scale

After adjustment for health status and other potential confounders the odds for specialist utilization in the last four weeks were still significantly higher under the EHC model (OR=2.39 [1.03-5.52]) as compared to the HV. Odds of GP utilization (OR= 2.18 [0.80-5.92]), specialist unmet needs (OR=2.01 [0.97-4.19]), emergency department use (OR= 1.35 [0.66-2.76]) and avoidable hospitalization (OR=1.77 [0.77-4.07]) were higher under the EHC model compared to the HV model, while there was nearly no difference for GP unmet needs. However, 95%-confidence intervals suggest that all differences are rather small and not or only marginally significant (c. Fig. 4).

**Figure 4:**
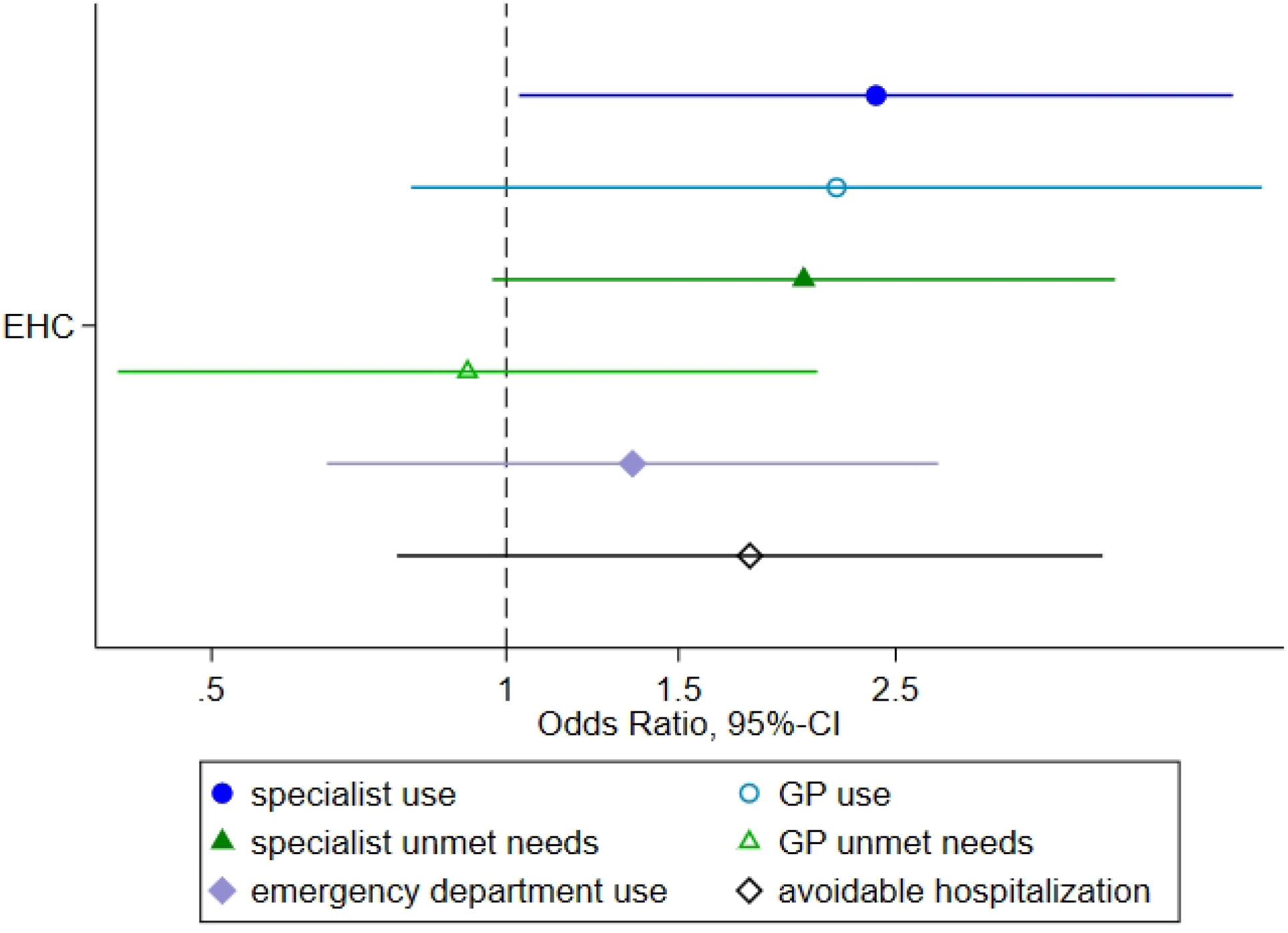
Fully adjusted Odds Ratios (and 95%-CIs) of access to healthcare comparing between access models used in the first 15 months (ref=HV) Legend: HV=Health care voucher; EHC=electronic health card; x-axis with 95% confidence intervals on a log-scale

Overall model-fit was acceptable with non-significant F-tests for nearly all final models (range of p-values between 0.420 and 0.849; p>0.001 for avoidable hospitalization). Analysis of the design effects for the final regression models (range of DEFF between 0.993 and 2.520) showed a moderate influence of weighting on the results, which stresses the importance of weighting for valid and generalizable results. However, the comparison of ORs between weighted and non-weighted results did not reveal major differences for any of the outcomes (see additional files 5-9).

## Discussion

Our study is the first comparison of realized access to healthcare between the three different access models for ASR in Germany. It thus adds important empirical knowledge to the current literature on access to healthcare among ASR. Our results show significant differences for specialist service utilization between the access models. ASR under the HV model reported lower needs-adjusted utilization of specialist services compared to persons using the EHC and to persons with regular access. Needs-adjusted GP utilization was also slightly but not significantly lower among people using HVs compared to both reference groups (EHC and regular access). Specialist unmet needs were also slightly lower among ASR with HV compared to ASR with EHC, but not compared to ASR with regular access. There was a slight tendency towards higher specialist unmet needs and avoidable hospitalizations among ASR with EHC compared to ASR with regular access, but no differences in the other outcomes.

The lower utilization of specialist services might be related to access barriers that are inherent to the HV model (such as the need for prior approval by the local welfare office for specialist utilization, or the limited validity of HVs to only three months). For all other outcomes – GP utilization, unmet needs, emergency department use and avoidable hospitalization – differences between the groups were neither consistent nor significant in the fully adjusted models. Tendencies towards differences in GP utilization, specialist unmet needs and avoidable hospitalizations should be further explored.

Using data from three population based, multi-lingual surveys with tested items resulted in high validity of the results. It also enabled us to control for a wide range of socio-demographic and health-related confounders inquired in the survey. Besides the immediate associations between access model and the outcomes, there are important methodological implications of our research. First, there was a significant association between region of origin and access model. This is mainly explained by the fact that the country of origin influences the distribution of newcomers among states in Germany as well as their chances of obtaining permanent legal status. At the same time, it highlights the importance of controlling for region or country of origin when making comparisons between the German states. Second, to the best of our knowledge, this was the first time that avoidable hospitalizations were assessed in a survey design (and not through routine or claims data using ICD-codes). The approach turned out to be feasible and the comparatively low share of missing responses (only 7.1%) showed a high acceptance among respondents. We thus consider the further assessment of the item’s validity and potential for use in future health surveys to be relevant. This is especially important for research on populations that tend to remain left out of routine data collection.

The study has some methodological limitations. While the questions referring to specialist and GP service utilization referred to the last four weeks, questions related to all other outcomes referred to the last 12 months. This may have led to a recall bias as people are requested to report their health seeking behaviour for long periods of time. In addition, respondents potentially changed from one access model to another (from HV or EHC to regular access), causing misclassification bias. Thus, results for unmet needs, emergency department use and avoidable hospitalization are less robust than for specialist and GP utilization. Observed minor differences for these outcomes (e.g., for specialist unmet needs) are therefore not further interpreted. Finally, our data is from two different states with potentially unmeasured differences in geographical accessibility, availability of interpreters and organization of healthcare services. These unmeasured differences might constitute confounders especially for the comparison between HV and EHC, as all included HV users lived in BW while all EHC users lived in BE.

Our results are in line with qualitative studies that hypothesised lower utilization of outpatient services among HV users due to bureaucratic barriers [19, 20]. A quantitative analysis of claims data [23] and a regional survey [22] in the state of North Rhine Westphalia, too, have identified access barriers related to specialist utilization, which ultimately led to inequalities in healthcare utilization. There is thus reason to suspect that persons who are subject to the HV model have lower access to specialist services compared to EHC users and people with regular access, while having equivalent needs. According to the literature on health inequalities this would constitute a violation of the principle of horizontal equity (equal access for equal needs) [39]. We did not find inequalities related to unmet needs, emergency department use and avoidable hospitalization. Other studies reported significant differences for these outcomes; for example, higher rates of avoidable hospitalizations among EHC users as compared to persons with regular access [32], and higher emergency department use under the HV model as compared to the EHC model [23]. We could not back up these findings with our analyses, which may be due to the abovementioned methodological limitations.

The results of our study have important implications for the controversial debate on the choice of access model for ASR during their first 18 months in Germany. The identified inequalities in access to specialist and GP services provide further evidence for the advantages of the EHC model compared to the HV model. The EHC model facilitates need-based healthcare utilization by providing access similar to the regular access model. Those local governments that, nonetheless, adhere to the HV model often justify their policy decision with cost arguments; that is, with the assumption that the EHC model would lead to excessive utilization of healthcare and thereby increase health expenses [40]. Given that recent studies refute such cost arguments [24, 41], little evidence-based arguments are left to justify upholding the HV model. In that light, policymakers who have so far opted for the HV model may want to reconsider introducing the EHC (or disclose the remaining reasons for not doing so). The relevance of such policy change has increased lately, as in August 2019 the waiting period during which the respective access models (HV and EHC) apply has been prolonged from 15 to 18 months [11].

While this study looked at the direct effects of the different models on access, we were unable to analyse long-term effects of lower healthcare utilization among HV users on their health status. Longitudinal studies will be needed to study the health consequences of the different access models. Such studies could revisit analyses of avoidable hospitalizations, emergency department use and unmet needs, as methodological limitations impeded a thorough analyses of these aspects in our study.

## Conclusion

Our comparison of the different access models for ASR shows that persons who are subject to the HV model are systematically disadvantaged in their access to healthcare. With equal need, they use specialist (and partly also GP) services less often than ASR with an EHC and those with regular access. The identified inequalities constitute inequities in access to healthcare that could be reduced by policy change from HV to the EHC model (or by granting regular access upon arrival). The EHC model ensures access to GP and specialist services comparable to regular access as there are no significant differences in outpatient care utilization between ASR with EHC and ASR with regular access. Interpretation of the results for unmet needs, emergency department use and avoidable hospitalization is limited due to methodological constrains. Still, the respective tendencies that were observed deserve further exploration in future studies.

## Data Availability

The datasets used during the current study are available from https://respond-study.org/en/resources/ on reasonable request to the corresponding author.

## List of Abbreviations

ACSC: Ambulatory care sensitive conditions
ASR: Asylum seekers and refugees
AsylbLG: Asylbewerberleistungsgesetz/Asylum Seeker’s Benefits Act
BW: Baden-Wuerttemberg
BE: Berlin
DEFF: Design effect
EHC: Electronic health card
GP: General practitioner
HV: Health care voucher
SWO: Social welfare office

## Declarations

### Ethics approval and consent to participate

The study has been positively reviewed by the Ethics Committee of the Charité University Hospital Berlin (EA4/111/18), and the ethics committee of the Medical Faculty Heidelberg (S-516/2017)

### Consent for publication

Not applicable

### Authors’ contributions

LB and KB conceived the study and were responsible for instrument development and sampling approach across both study sites, as well as data collection and methodology in Baden-Württemberg. NG was responsible for data collection and methodology in Berlin. JW and KB conceptualized the analysis plan. JW conducted the analysis and drafted the first version of the manuscript. LB was responsible for data management and contributed significantly to the formal analysis. LB, NG and KB revised the manuscript critically for important intellectual content. All authors read and approved the final manuscript.

## Acknowledgements

The authors would like to thank all study participants for their valuable time and efforts.

LB and KB would like to thank the Association of Districts (Landkreistag) of Baden-Württemberg, the district authorities all 44 districts responsible for reception of ASR, the Ministry of Social Affairs and Integration Baden-Württemberg, the Ministry of the Interior, Digitalization and Migration Baden-Württemberg, and the responsible regional councils of the state of Baden-Württemberg for supporting the study.

NG would like to thank those students of the Berlin School of Public Health, who were involved in the Berlin-based data collection for this study; and she is indebted to the Berlin Senate Administration, the Berlin Office for Refugee Affairs, and the managers and staff of the included accommodation centres for supporting this research.

## List of additional files

Additional file 1 (pdf): Participants’ flow diagram and response rate

Additional file 2 (pdf): List of ambulatory care-sensitive condition

Additional file 3 (pdf): Overview of clusters, stratification, and survey weights

Additional file 4 (pdf): Overview of imputation models

Additional file 5 (pdf): Detailed results related to Figure 1 – Results of logistic regression model (odds ratios and standard errors)

Additional file 6 (pdf): Detailed results related to Figure 2 – Results of logistic regression model (odds ratios and standard errors)

Additional file 7 (pdf): Detailed results related to Figure 3 – Results of logistic regression model (odds ratios and standard errors)

Additional file 8: Detailed results related to Figure 4 – Results of logistic regression model (odds ratios and standard errors)

Additional file 9: Overview of effect of weighting on main outcomes in final regression models (design effects/DEFF)

