## Supplementary figures and images for "Inequalities in access to healthcare by local policy model among newly arrived refugees: evidence from population-based studies in two German states"

### Additional file 1

#### Additional file 1: Participants' flow diagram and response rate

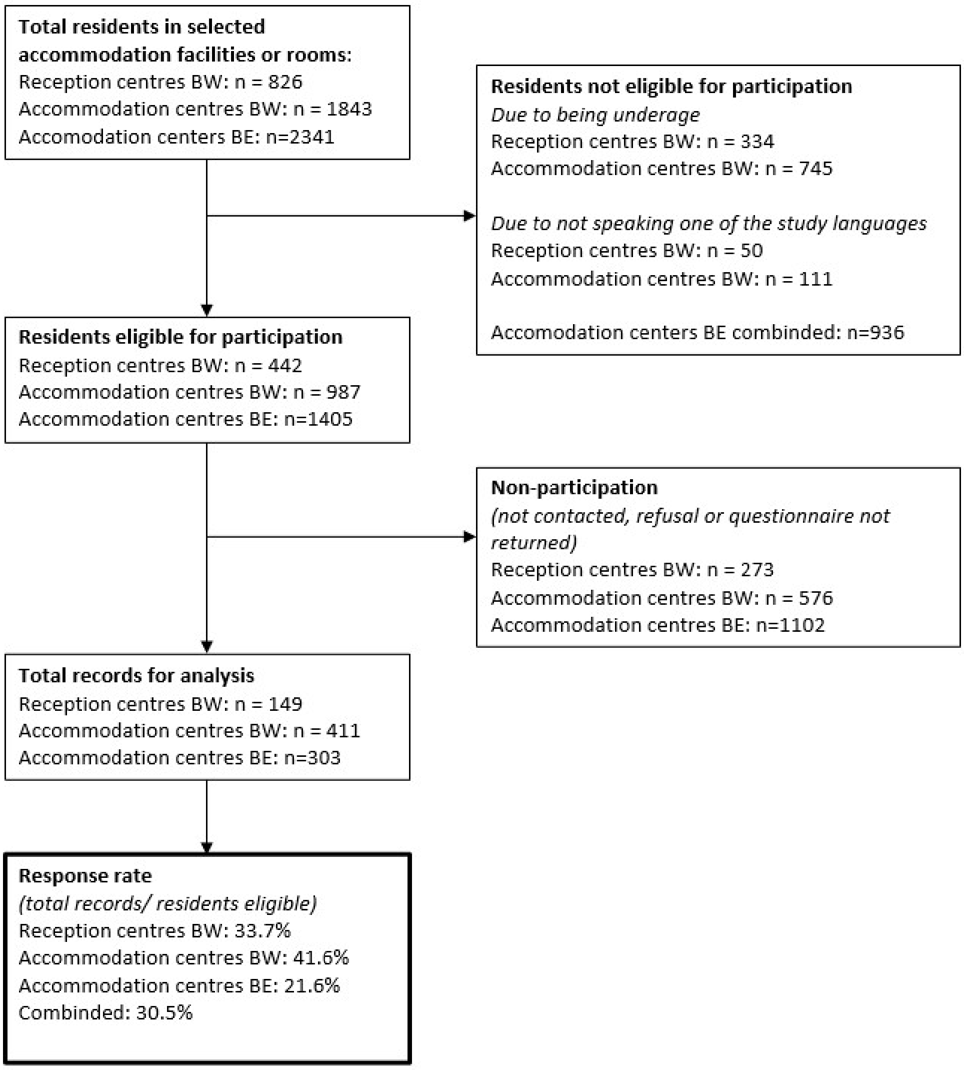
