## Additional file 2 for "Inequalities in access to healthcare by local policy model among newly arrived refugees: evidence from population-based studies in two German states"

### Additional file 2: List of ambulatory care-sensitive condition

| |
|---|
| Stroke |
| Angina |
| Heart failure |
| High blood pressure |
| Bronchitis |
| Mental or behavioural disorders due to substance abuse |
| Depression |
| Back pain |
| Diarrhoea |
| Flu |
| Ear, nose, and throat infections |
| Diabetes |
| Epilepsy |
| Sleeping problems |
| Cavities |
