## Additional file 3 for "Inequalities in access to healthcare by local policy model among newly arrived refugees: evidence from population-based studies in two German states"

**Additional file 3: Overview of clusters, stratification, and survey weights**

| Level | ACs BW | RCs BW | ACs BE | Cluster | Stratification (size) | Finite population correction |
|---|---|---|---|---|---|---|
| Level 1 | We randomly selected several ACs from all possible units (prob. of inclusion ca. 3%) | We deterministically chose several RCs based on location and size (prob. of inclusion 100%) | We randomly selected several ACs from all possible units (prob. of inclusion ca. 3%) | AC/RC: facility | *Strata1 (BW)*<br>AC: 33<br>RC: 1<br>*Strata2 (BW+BE)*<br>AC-BW: 19<br>RC-BW: 1<br>AC-BE 3 | Total no. of AC:<br>BW n=1945<br>BE n=72 |
| Level 2 | We attempted to recruit all **individuals** within each facility (prob. of inclusion 100%) | We randomly chose a number of **rooms** in each facility (prob. of inclusion ca. 25%) | We attempted to recruit all **individuals** within each facility (prob. of inclusion 100%) | AC: individuals<br>RC: rooms | | Total no. of rooms/individuals in chosen facility (n=1123) |

AC=accommodation centre; RC=reception centre, BW=Baden-Wuerttemberg; BE=Berlin
