## Additional file 4 for "Inequalities in access to healthcare by local policy model among newly arrived refugees: evidence from population-based studies in two German states"

### Additional file 4: Overview of imputation models

| Model variables for imputation | Imputation method |
|---|---|
| Age | Predictive mean matching |
| sex | Logistic regression model |
| Nationality (region of origin) | Polytomous regression model |
| Educational score | Predictive mean matching |
| Residence status | Polytomous regression model |
| Time since arrival (months) | Predictive mean matching |
| Access model | Polytomous regression model |
| GP unmet needs | Logistic regression model |
| Specialist unmet needs | Logistic regression model |
| GP 4-week-utilization | Logistic regression model |
| Specialist 4-week-utilization | Logistic regression model |
| Emergency department use | Logistic regression model |
| Avoidable hospitalization | Logistic regression model |
| Chronical illness | Logistic regression model |
| General health | Polytomous regression model |
| Family doctor | Logistic regression model |
