## Additional file 5 for "Inequalities in access to healthcare by local policy model among newly arrived refugees: evidence from population-based studies in two German states"

**Additional file 5: Detailed results related to Figure 1 – Results of logistic regression model (odds ratios and standard errors)**

| | *Specialist use* | *GP use* | *Specialist unmet needs* | *GP unmet needs* | *Emergency dept. use* | *Avoidable hospitalization* |
|---|---|---|---|---|---|---|
| *HV (ref. regular access)* | 0.466*** | 0.571** | 1.095 | 0.915 | 0.761 | 0.996 |
| | (0.0937) | (0.145) | (0.256) | (0.214) | (0.152) | (0.230) |
| *EHC (ref. regular access)* | 0.899 | 0.814 | 2.119*** | 1.050 | 1.034 | 1.514 |
| | (0.279) | (0.339) | (0.496) | (0.304) | (0.330) | (0.465) |
| *Age* | 1.015* | 1.028*** | 1.018* | 1.012 | 0.995 | 1.015 |
| | (0.00852) | (0.0101) | (0.00896) | (0.00948) | (0.00918) | (0.00965) |
| *Male (ref. female)* | 1.507** | 1.719*** | 1.106 | 1.106 | 2.319*** | 1.145 |
| | (0.270) | (0.334) | (0.207) | (0.218) | (0.419) | (0.254) |
| *Constant* | 0.466*** | 0.571** | 1.095 | 0.915 | 0.761 | 0.996 |
| | (0.0937) | (0.145) | (0.256) | (0.214) | (0.152) | (0.230) |
| *Observations* | 863 | 863 | 863 | 863 | 863 | 863 |
| *p-value (F-test)* | 0.990 | 0.708 | 0.567 | 0.945 | 0.083 | 0.191 |

*Standard errors in parentheses; *** p<0.01, ** p<0.05, * p<0.1*
