## Additional file 6 for "Inequalities in access to healthcare by local policy model among newly arrived refugees: evidence from population-based studies in two German states"

**Additional file 6: Detailed results related to Figure 2 – Results of logistic regression model (odds ratios and standard errors)**

| | *Specialist use* | *GP use* | *Specialist unmet needs* | *GP unmet needs* | *Emergency dept. use* | *Avoidable hospitalization* |
|---|---|---|---|---|---|---|
| *HV (ref. regular access)* | 0.405*** | 0.614 | 0.937 | 0.953 | 0.736 | 0.952 |
| | (0.0972) | (0.192) | (0.243) | (0.258) | (0.158) | (0.291) |
| *EHC (ref. regular access)* | 0.967 | 1.336 | 1.887* | 0.870 | 0.991 | 1.689 |
| | (0.360) | (0.593) | (0.612) | (0.314) | (0.321) | (0.558) |
| *Age* | 1.001 | 0.999 | 1.004 | 0.990 | 0.976** | 0.991 |
| | (0.00916) | (0.00992) | (0.0101) | (0.0109) | (0.0117) | (0.0116) |
| *Male (ref. female)* | 1.386* | 1.619** | 1.054 | 1.039 | 2.238*** | 1.035 |
| | (0.255) | (0.344) | (0.205) | (0.228) | (0.413) | (0.262) |
| *Time since arrival (months)* | 0.997 | 1.004 | 0.999 | 1.006 | 0.995 | 0.998 |
| | (0.00351) | (0.00401) | (0.00307) | (0.00367) | (0.00332) | (0.00356) |
| *Region: Asia (ref. Europe)* | 1.000 | 1.880** | 1.246 | 0.928 | 0.750 | 1.026 |
| | (0.307) | (0.540) | (0.441) | (0.286) | (0.184) | (0.290) |
| *Region: Africa (ref. Europe)* | 1.358 | 1.571 | 1.232 | 0.949 | 0.910 | 1.563 |
| | (0.515) | (0.538) | (0.550) | (0.334) | (0.332) | (0.631) |
| *Region: Other (ref. Europe)* | 1.330 | 1.991* | 1.180 | 1.015 | 1.229 | 1.312 |
| | (0.618) | (0.765) | (0.475) | (0.444) | (0.520) | (0.655) |

| | | | | | | |
|---|---|---|---|---|---|---|
| *Rather bad general health (ref. rather good)* | 2.430*** | 1.854** | 2.323*** | 2.363*** | 1.499* | 1.752** |
| | (0.478) | (0.504) | (0.437) | (0.576) | (0.345) | (0.454) |
| *No chronical illness (ref. yes)* | 1.099 | 2.516*** | 2.069*** | 2.482*** | 1.789** | 3.103*** |
| | (0.223) | (0.681) | (0.458) | (0.600) | (0.436) | (0.784) |
| *Education: low (ref. high)* | 1.318 | 0.868 | 1.009 | 0.644* | 1.133 | 1.233 |
| | (0.332) | (0.201) | (0.246) | (0.152) | (0.320) | (0.325) |
| *Education: middle (ref. high)* | 1.412 | 0.933 | 0.907 | 0.821 | 0.763 | 0.749 |
| | (0.317) | (0.243) | (0.195) | (0.195) | (0.203) | (0.206) |
| *No family doctor (ref. yes)* | 1.931*** | 3.209*** | 0.643** | 0.922 | 1.886*** | 1.798*** |
| | (0.451) | (0.663) | (0.120) | (0.186) | (0.414) | (0.337) |
| *Reception centre (ref. accommodation centre)* | 1.498 | 1.975** | 0.776 | 0.819 | 0.870 | 0.755 |
| | (0.415) | (0.606) | (0.336) | (0.305) | (0.244) | (0.269) |
| *Constant* | 0.166*** | 0.100*** | 0.300 | 0.436 | 0.766 | 0.179** |
| | (0.105) | (0.0657) | (0.224) | (0.302) | (0.467) | (0.124) |
| *Observations* | 863 | 863 | 863 | 863 | 863 | 863 |
| *p-value (F-test)* | 0.479 | 0.779 | 0.420 | 0.681 | 0.849 | >0.001 |

*Standard errors in parentheses; *** p<0.01, ** p<0.05, * p<0.1*
