## Additional file 7 for "Inequalities in access to healthcare by local policy model among newly arrived refugees: evidence from population-based studies in two German states"

**Additional file 7: Detailed results related to Figure 3 – Results of logistic regression model (odds ratios and standard errors)**

| | *Specialist use* | *GP use* | *Specialist unmet needs* | *GP unmet needs* | *Emergency dept. use* | *Avoidable hospitalization* |
|---|---|---|---|---|---|---|
| *EHC (ref. HV)* | 1.930** | 1.425 | 1.935** | 1.147 | 1.359 | 1.521 |
| | (0.618) | (0.673) | (0.513) | (0.360) | (0.495) | (0.536) |
| *Age* | 1.015* | 1.028*** | 1.018* | 1.012 | 0.995 | 1.015 |
| | (0.00852) | (0.0101) | (0.00896) | (0.00948) | (0.00918) | (0.00965) |
| *Male (ref. female)* | 1.507** | 1.719*** | 1.106 | 1.106 | 2.319*** | 1.145 |
| | (0.270) | (0.334) | (0.207) | (0.218) | (0.419) | (0.254) |
| *Constant* | 0.217*** | 0.309*** | 0.288*** | 0.315*** | 0.419** | 0.204*** |
| | (0.0697) | (0.104) | (0.0949) | (0.0951) | (0.142) | (0.0598) |
| *Observations* | 863 | 863 | 863 | 863 | 863 | 863 |
| *p-value (F-test)* | 0.990 | 0.708 | 0.567 | 0.945 | 0.083 | 0.191 |

*Standard errors in parentheses; *** p<0.01, ** p<0.05, * p<0.1*
