## Additional file 9 for "Inequalities in access to healthcare by local policy model among newly arrived refugees: evidence from population-based studies in two German states"

**Additional file 9: Overview of effect of weighting on main outcomes in final regression models (design effects/DEFF)**

| | *Specialist use* | *GP use* | *Specialist unmet needs* | *GP unmet needs* | *Emergency dept. use* | *Avoidable hospitalization* |
|---|---|---|---|---|---|---|
| *HV (ref. regular access)* | 1.485 | 2.520 | 1.792 | 1.911 | 1.260 | 1.929 |
| *EHC (ref. regular access)* | 1.216 | 1.596 | 1..137 | 1.172 | 1.032 | 0.993 |
| *EHC (ref. HV)* | 1.257 | 1.746 | 1.174 | 1.259 | 1.038 | 1.229 |
